# High-Quality Masks Reduce COVID-19 Infections and Deaths in the US

**DOI:** 10.1101/2020.09.27.20199737

**Authors:** Erik Rosenstrom, Buse Eylul Oruc, Nathaniel Hupert, Julie Ivy, Pinar Keskinocak, Maria E. Mayorga, Julie L. Swann

## Abstract

**Objectives:** To evaluate the effectiveness of widespread adoption of masks or face coverings to reduce community transmission of the SARS-CoV-2 virus that causes COVID-19.

**Methods:** We created an agent-based stochastic network simulation using a variant of the standard SEIR dynamic infectious disease model. We considered a mask order that was initiated 3.5 months after the first confirmed COVID-19 case. We varied the likelihood of individuals wearing masks from 0-100% in steps of 20% (mask adherence) and considered 25% to 90% mask-related reduction in viral transmission (mask efficacy). Sensitivity analyses assessed early (by week 13) versus late (by week 42) adoption of masks and geographic differences in adherence (highest in urban and lowest in rural areas).

**Results:** Introduction of mask use with 50% efficacy worn by 50% of individuals reduces the cumulative infection attack rate (IAR) by 27%, the peak prevalence by 49%, and population-wide mortality by 29%. If 90% of individuals wear 50% efficacious masks, this decreases IAR by 54%, peak prevalence by 75%, and population-wide mortality by 55%; similar improvements hold if 70% of individuals wear 75% efficacious masks. Late adoption reduces IAR and deaths by 18% or more compared to no adoption. Lower adoption in rural areas than urban would lead to rural areas having the highest IAR.

**Conclusions:** Even after community transmission of SARS-CoV-2 has been established, adoption of mask-wearing by a majority of community-dwelling individuals can meaningfully reduce the number and outcome of COVID-19 infections over and above physical distancing interventions.

**Highlights:**

- This paper shows the impact of widespread adoption of masks in response to the COVID-19 pandemic, with varying levels of population adherence, mask efficacy, and timing of mask adoption.
- The paper’s findings help inform messaging to policymakers at the state or local level considering adding or keeping mask mandates, and to communities to promote widespread adoption of high-quality masks.
- Adoption of masks by at least half of the population can reduce cumulative infections and population deaths by more than 25%, while decreasing peak prevalence by about 50%. Even greater marginal improvements arise with adoption rates above 70%. The benefits of adopting high-quality masks is above that achieved by mobility changes and distancing alone.
- Rural and suburban areas are at higher relative risk than urban areas, due to less distancing and lower adoption of masks.

## Introduction

Since its introduction in early 2020, the SARS-CoV-2 coronavirus that causes COVID-19 has been widely circulating throughout the United States [1]. While there is growing empirical evidence that masks can be effective at reducing SARS-CoV-2 transmission via droplets [2, 3], the impact of population-based mask use on the COVID-19 pandemic is still poorly defined. Some states were early mask promoters (e.g., North Carolina or NC in June), some were late (e.g., North Dakota in November), and some have remained silent (e.g., South Dakota). This study prospectively assesses the effectiveness of face coverings at the state level and across urban, suburban, and rural counties under different mask efficacy and population adherence levels while providing additional evidence for the benefit of existing adoptions. We focus on masks because they are relatively inexpensive and logistically non-disruptive (compared to, e.g., physical distancing measures). Masks are also a fast intervention that can be adopted if SARS-Cov-2 mutates or if other viruses arise. We include analysis of state-wide effects along with those on subpopulations stratified by geography and demographics.

## METHODS

We employ an agent-based stochastic network model with an SEIR framework [4] for the progression of SARS-CoV-2 [5, 6]. As in Keskinocak et al [6], we simulate interactions among agents, where transmission can occur daily in households, workplaces and schools, and community settings, with day/night differentiation in interactions. The model is also similar to Patel et al [7], where we study vaccine distribution. The network of agents is built using US Census data [8] at the tract level using household size, presence of children, and age groups (defined as <= 4,5-9, 10-19, 20-64, or 65 years old and above). Model parameters include the reproductive rate (2.4 without interventions), hospitalization and mortality rates (by age group), an effective overall Infection Fatality Rate just under 0.5%, and asymptomatic and symptomatic transmission coefficients [5-7]. We also incorporate race/ethnicity in the model at the household level, as well as the existence of a COVID-19-associated health condition (diabetes) stratified by race/ethnicity using available data [9]. Full details are outlined in Supplementary Materials available online.

Analysis was performed for the representative state of North Carolina (NC), which has had SARS-CoV-2 transmission rate that puts it in the lower half of US states as quantified by deaths per capita [1]. The population of 10.5 million people is represented with a sample of 1,017,720 agents. The simulation is seeded (day 1) with cumulative cases as of March 24, 2020 [1], where the initial cases are multiplied by 10 to account for underreporting [10] and scaled to the number of agents in the simulation; the simulation is run for 365 days. County cases are distributed to households randomly across each county’s census tracts proportional to the tract population. We categorize each census tract by urban/suburban/rural status, where urban corresponds to Rural-Urban-Commuting-Area (RUCA) codes of 1,2 for urban; 6,7,8,9,10 for rural; and the remainder for suburban [11].

The model captures the likelihood an adult agent stays home over time using SafeGraph data [12] (see supplement for details) aggregated by month and census tract. SafeGraph data captures the presence of devices in homes or other settings over time and across census blocks, which we aggregated into tracts grouped by urbanicity (urban, rural, suburban) and median household income (4 quartiles statewide). An adult will work from home on a given day according to a probability drawn from their census tract’s rate. For interactions with the community by age group, we assume the rate follows the same pattern as the workplace mobility data but with a smaller reduction compared to workplace (e.g., a 40% reduction in work attendance is associated with a 12% reduction in community interaction), see Supplemental Appendix). This is consistent with our comparisons of workplace mobility with other types of mobility [12, 13]. We assume mobility rates stabilize at month six levels. We do not assume a link between mobility and population infections as we did not see it consistently across locations when comparing mobility data and infections. The mobility data captures the fact that many people stayed home shortly after cases began rising (consistent with shelter-at-home orders given in NC and many other states) and mobility continues to be lower than pre-pandemic rates. We assume that households with a symptomatic Covid-19 infection will voluntarily quarantine (VQ), in line with the low quarantining rates from [6] (see Supplemental Appendix). Schools are virtual or closed initially, opening on month six with students rotating every other day. Anyone who is symptomatic stays home from school and away from work peer groups.

Unlike Keskinocak et al [6], a proportion of the population wears masks under early and late scenarios. In the early scenario, the adherence rate increases approximately monthly (days 6-94, where the last date corresponds to the state mask order in NC), linearly from 0 to the final adherence probability of (0, 40, 60, 80, or 100%). The “late” scenario, where mask adoption reaches the highest level when the cumulative infections are approximately equal to 15% in the baseline cases (specifically, on day 282), corresponds to later mask orders such as the one issued in North Dakota [14]. In the baseline scenarios, we assume the mask adherence is homogeneous across the population, as supported by large-scale random sampling of the population from July 2020 [15]. In sensitivity analysis, we allow mask adherence to vary by urbanicity, [85%,75%,65%] or [85%,70%,55%] based on the November 2020 surveys conducted by Facebook [16], with values representing typical ones for states with higher or lower mask usage (e.g., NC that had early adoption vs ND that had late adoption). Based on recent experimental analysis [3], in the baseline scenarios we assume that masks reduce the infectivity to others and susceptibility by 50% each (mask efficacy); if two agents encounter each other in the simulation, the potential for encounter-based transmission reduction is multiplicative if both are wearing masks. We compare the baseline mask efficacy with scenarios where higher quality, more efficacious masks are employed (e.g., 80+% reduction in transmission and susceptibility risk, as may be achieved by surgical, N95, or even multi-layer sewn masks). Note that the population-level effectiveness of masks can be computed as adherence multiplied by efficacy, e.g., 75% adherence times 50% efficacy equals 37.5% effectiveness. For our no-intervention control, we assume there are no interventions throughout the pandemic and mobility is as normal; there is also a scenario with 0% mask adherence that has changes in mobility. Note that we do not assume a direct relationship between rising infections and other aspects such as distancing (captured by mobility), mask usage, or changes in fatality rates.

The model is validated against reported hospitalizations and deaths in NC as of 11/1/2020, where the validation accounts for the fact that not all positive cases are lab-reported (See the Supplemental Appendix). All simulation output values are adjusted to the true population of 10.49 million.

We compute the infection attack rate (IAR) of all infections over the time horizon, the peak percentage of the population simultaneously infected, the peak count of hospitalizations, and the mortality of the total population (total deaths or a mortality rate for subpopulations. Since the model is stochastic, we quantify the mean and standard deviation values over 15 replications, where the number of replications balances reductions in variability of measures such as IAR and total deaths with computational time. We provide values at the state level, by county, and stratified by urban/suburban/rural status. In the studied state, 75% of people are in urban areas, 15% in suburban, and 10% in rural areas. We use the phrase percentage points if referring to absolute differences or give a percentage if the change is relative (e.g., a drop from 30% to 20% IAR is 10 percentage points or a 33% reduction).

The funding supported the students performing the analysis; it had no relationship with the design of the experiments, interpretation of the results, or determining the conclusions.

This study uses publicly available de-identified data and does not require IRB approval.

## RESULTS

Even at low levels of effectiveness, mask-wearing reduces cumulative infections, peak infections, hospitalizations, and mortality for COVID-19 (Table 1). If 75% of individuals wear 50% efficacious masks (population-level effectiveness of 37.5%), the IAR, peak infection, and deaths are reduced by 37%, 68%, and 47%, respectively, vs. no mask use, even in the setting of effective physical distancing measures..

**Table 1:**
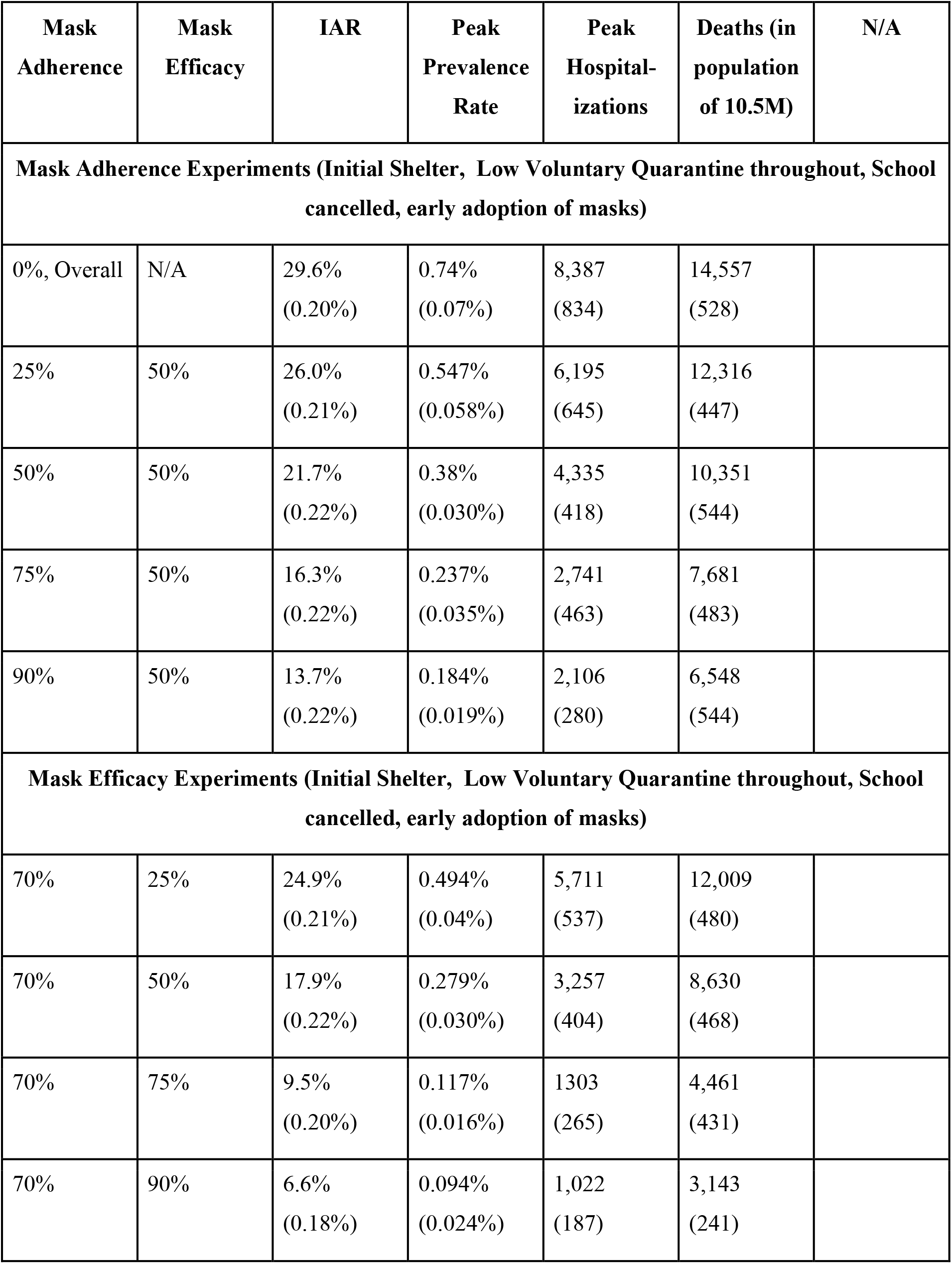
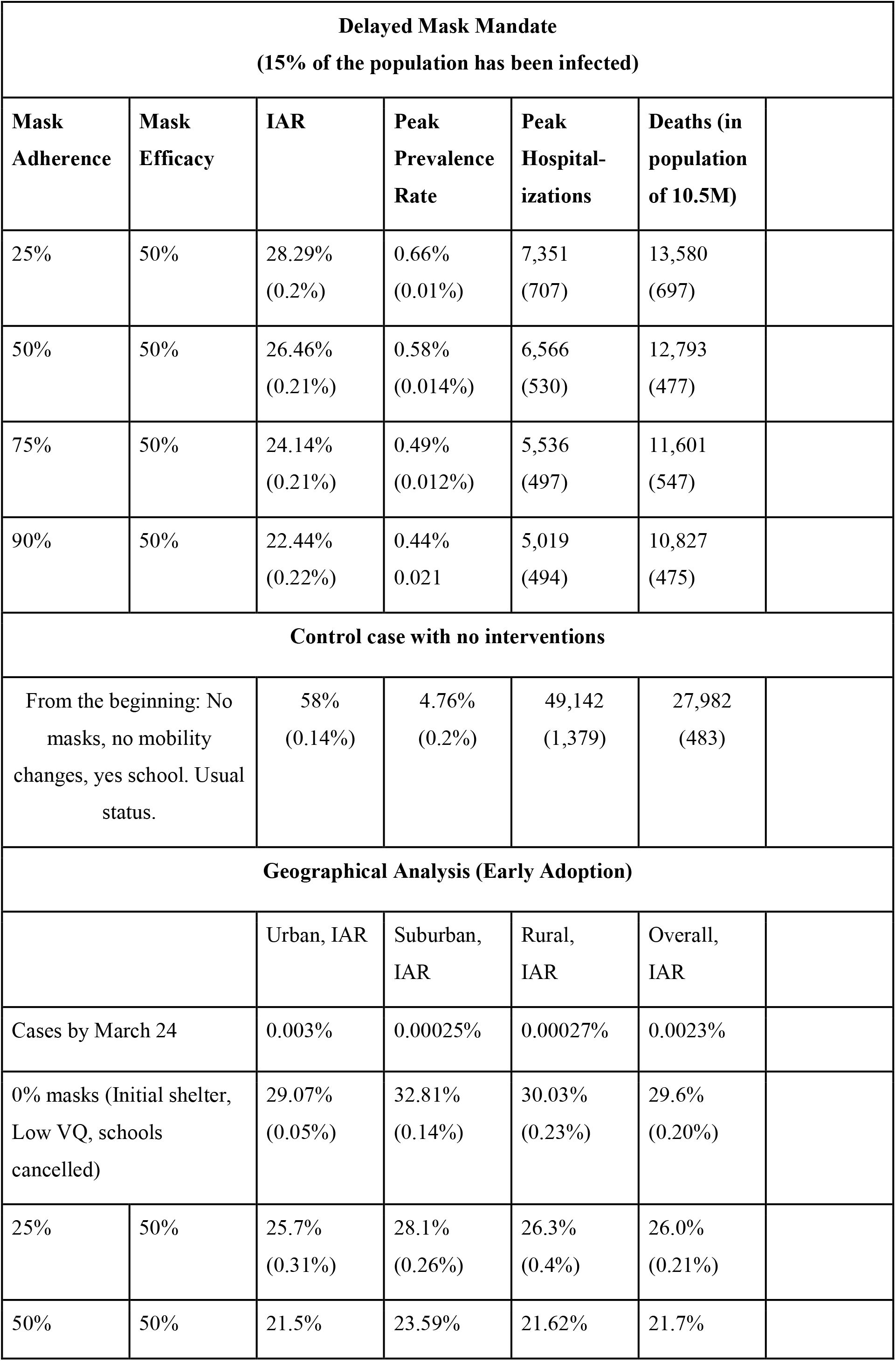
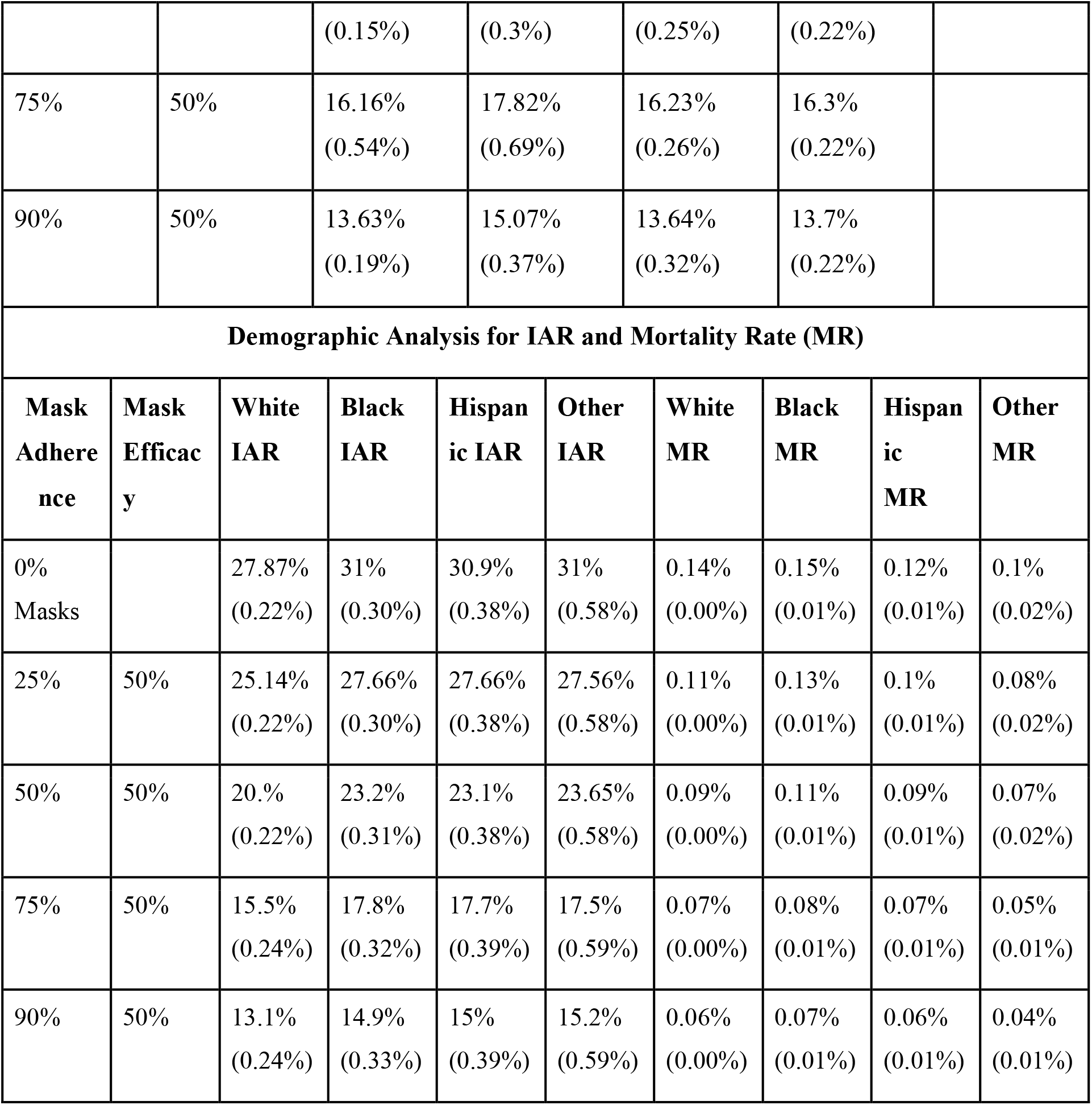
The results of the mask adherence and efficacy scenarios are shown for the metrics of IAR, peak prevalence rate, peak hospitalizations, and deaths for a state population of 10.5 million with mean (stdev) displayed. Highlighted rows are used in figure comparisons.

| Mask Adherence | Mask Efficacy | IAR | Peak Prevalence Rate | Peak Hospitalizations | Deaths (in population of 10.5M) | N/A |
| --- | --- | --- | --- | --- | --- | --- |
| **Mask Adherence Experiments (Initial Shelter, Low Voluntary Quarantine throughout, School cancelled, early adoption of masks)** |  |  |  |  |  |  |
| 0%, Overall | N/A | 29.6%<br>(0.20%) | 0.74%<br>(0.07%) | 8,387<br>(834) | 14,557<br>(528) |  |
| 25% | 50% | 26.0%<br>(0.21%) | 0.547%<br>(0.058%) | 6,195<br>(645) | 12,316<br>(447) |  |
| 50% | 50% | 21.7%<br>(0.22%) | 0.38%<br>(0.030%) | 4,335<br>(418) | 10,351<br>(544) |  |
| 75% | 50% | 16.3%<br>(0.22%) | 0.237%<br>(0.035%) | 2,741<br>(463) | 7,681<br>(483) |  |
| 90% | 50% | 13.7%<br>(0.22%) | 0.184%<br>(0.019%) | 2,106<br>(280) | 6,548<br>(544) |  |
| **Mask Efficacy Experiments (Initial Shelter, Low Voluntary Quarantine throughout, School cancelled, early adoption of masks)** |  |  |  |  |  |  |
| 70% | 25% | 24.9%<br>(0.21%) | 0.494%<br>(0.04%) | 5,711<br>(537) | 12,009<br>(480) |  |
| 70% | 50% | 17.9%<br>(0.22%) | 0.279%<br>(0.030%) | 3,257<br>(404) | 8,630<br>(468) |  |
| 70% | 75% | 9.5%<br>(0.20%) | 0.117%<br>(0.016%) | 1303<br>(265) | 4,461<br>(431) |  |
| 70% | 90% | 6.6%<br>(0.18%) | 0.094%<br>(0.024%) | 1,022<br>(187) | 3,143<br>(241) |  |

| **Delayed Mask Mandate**<br>**(15% of the population has been infected)** |  |  |  |  |  |
| --- | --- | --- | --- | --- | --- |
| Mask Adherence | Mask Efficacy | IAR | Peak Prevalence Rate | Peak Hospitalizations | Deaths (in population of 10.5M) |
| 25% | 50% | 28.29%<br>(0.2%) | 0.66%<br>(0.01%) | 7,351<br>(707) | 13,580<br>(697) |
| 50% | 50% | 26.46%<br>(0.21%) | 0.58%<br>(0.014%) | 6,566<br>(530) | 12,793<br>(477) |
| 75% | 50% | 24.14%<br>(0.21%) | 0.49%<br>(0.012%) | 5,536<br>(497) | 11,601<br>(547) |
| 90% | 50% | 22.44%<br>(0.22%) | 0.44%<br>0.021 | 5,019<br>(494) | 10,827<br>(475) |
| Control case with no interventions |  |  |  |  |  |
| From the beginning: No masks, no mobility changes, yes school. Usual status. |  | 58%<br>(0.14%) | 4.76%<br>(0.2%) | 49,142<br>(1,379) | 27,982<br>(483) |
| Geographical Analysis (Early Adoption) |  |  |  |  |  |
|  |  | Urban, IAR | Suburban, IAR | Rural, IAR | Overall, IAR |
| Cases by March 24 |  | 0.003% | 0.00025% | 0.00027% | 0.0023% |
| 0% masks (Initial shelter, Low VQ, schools cancelled) |  | 29.07%<br>(0.05%) | 32.81%<br>(0.14%) | 30.03%<br>(0.23%) | 29.6%<br>(0.20%) |
| 25% | 50% | 25.7%<br>(0.31%) | 28.1%<br>(0.26%) | 26.3%<br>(0.4%) | 26.0%<br>(0.21%) |
| 50% | 50% | 21.5% | 23.59% | 21.62% | 21.7% |

|  |  |  |  |  |  |  |  |  |  |
| --- | --- | --- | --- | --- | --- | --- | --- | --- | --- |
|  |  | (0.15%) | (0.3%) | (0.25%) | (0.22%) |  |  |  |  |
| 75% | 50% | 16.16%<br>(0.54%) | 17.82%<br>(0.69%) | 16.23%<br>(0.26%) | 16.3%<br>(0.22%) |  |  |  |  |
| 90% | 50% | 13.63%<br>(0.19%) | 15.07%<br>(0.37%) | 13.64%<br>(0.32%) | 13.7%<br>(0.22%) |  |  |  |  |
| Demographic Analysis for IAR and Mortality Rate (MR) |  |  |  |  |  |  |  |  |  |
| Mask Adherence | Mask Efficacy | White IAR | Black IAR | Hispanic IAR | Other IAR | White MR | Black MR | Hispanic MR | Other MR |
| 0% Masks |  | 27.87%<br>(0.22%) | 31%<br>(0.30%) | 30.9%<br>(0.38%) | 31%<br>(0.58%) | 0.14%<br>(0.00%) | 0.15%<br>(0.01%) | 0.12%<br>(0.01%) | 0.1%<br>(0.02%) |
| 25% | 50% | 25.14%<br>(0.22%) | 27.66%<br>(0.30%) | 27.66%<br>(0.38%) | 27.56%<br>(0.58%) | 0.11%<br>(0.00%) | 0.13%<br>(0.01%) | 0.1%<br>(0.01%) | 0.08%<br>(0.02%) |
| 50% | 50% | 20.%<br>(0.22%) | 23.2%<br>(0.31%) | 23.1%<br>(0.38%) | 23.65%<br>(0.58%) | 0.09%<br>(0.00%) | 0.11%<br>(0.01%) | 0.09%<br>(0.01%) | 0.07%<br>(0.02%) |
| 75% | 50% | 15.5%<br>(0.24%) | 17.8%<br>(0.32%) | 17.7%<br>(0.39%) | 17.5%<br>(0.59%) | 0.07%<br>(0.00%) | 0.08%<br>(0.01%) | 0.07%<br>(0.01%) | 0.05%<br>(0.01%) |
| 90% | 50% | 13.1%<br>(0.24%) | 14.9%<br>(0.33%) | 15%<br>(0.39%) | 15.2%<br>(0.59%) | 0.06%<br>(0.00%) | 0.07%<br>(0.01%) | 0.06%<br>(0.01%) | 0.04%<br>(0.01%) |

Higher mask adherence leads to improvement in each metric, and the improvement is not necessarily linear. Increasing adherence from 50% to 75% has a higher incremental improvement in IAR points (5.4) than increasing from 0 to 25% or 25% to 50% (3.6 and 4.3, respectively). A similar improvement is seen in the number of deaths, which drops by 25.8% when adherence increases from 50% to 75%.

Notably, in the best scenario we studied where 90% of people wear a mask that is 50% efficacious, this results in an almost 50% reduction in IAR to 13.7% (compared to 21.7% with 50% adherence); additionally, peak hospitalizations decrease by 51% and deaths by 36.7% in this scenario.

If higher quality masks are worn, then all metrics improve. As with mask adherence, the incremental improvement is nonlinear. For example, the IAR decreases incrementally by 7, 8.4, and 2.9 percentage points over the previous value as mask efficacy increases (from 25% to 50%, then to 75%, then to 90%, respectively, all with adherence of 70%). The incremental reduction in the number of deaths is also highest as efficacy increases from 50 to 75%.

Performance measures are better if mask adherence is adopted early (starting week 1), but late mask adherence (starting week 40) still improves population-based outcomes (see Figure 1, which shows the prevalence over time for mask adherence, efficacy, and timing, and Table 1 for corresponding summary values). Early adoption has a cumulative IAR that is 4.8 to 8.7 percentage points better than the corresponding late adoption case, depending on other parameters. However, even late adoption of mask use with adherence levels of 50%-90% improves the cumulative IAR over the no mask case by 3.1-7.2 percentage points, and reduces the number of deaths by 12.1%-25.6%. Late adoption with high mask adherence (90%) is marginally better than early adoption with low adherence (25% or 50%).

**Figure 1:**
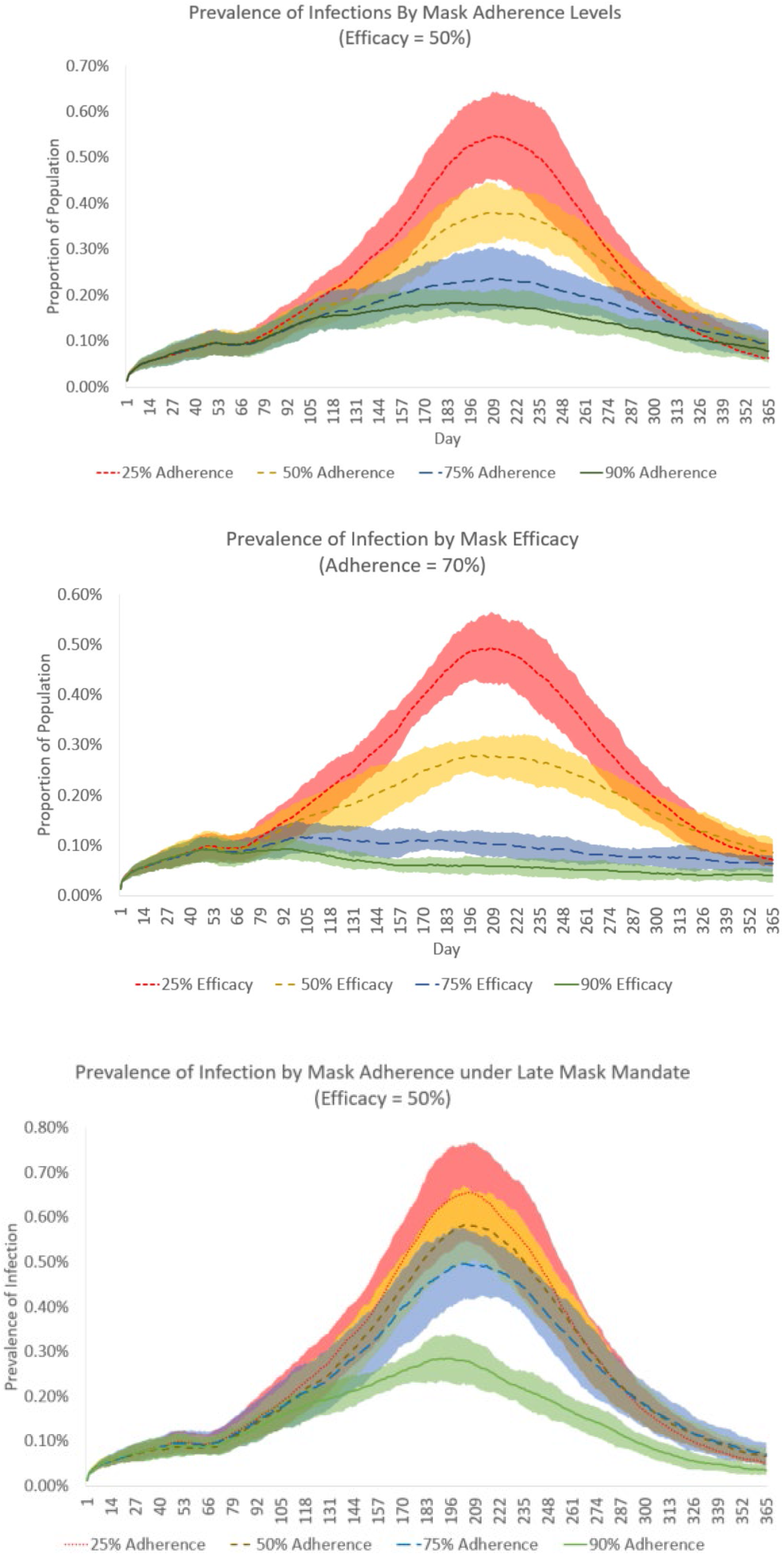
Prevalence of infectious people over time is shown. Subfigure (top) shows prevalence of cases with different levels of population adherence (25%, 50%, 75%, 90%) while mask efficacy is 50%. Subfigure (middle) shows prevalence with different levels of mask efficacy (25%, 50%, 75%, 90) while population adherence is 70%. Subfigure (bottom) shows prevalence with different levels of population adherence under late adoption of masks.

The results vary somewhat by RUCA county type, see Table 1 and Figure 2. Note that mobility differences across area types (shown in the Supplemental Appendix) indicate that urban areas have stayed home at higher rates than other areas, d with rural areas remaining the most mobile. In the baseline scenario (50% adherence): the IAR for suburban areas is about two points higher than that of urban and rural areas (23.59% versus 21.5% and 21.62%). The difference between urban and rural areas is biggest with 0% mask adherence (25.7% versus 26.3%) and smallest with 90% adherence (13.63% and 13.64%).When mask adherence differs across geographies as seems indicated in recent data, then the IAR tends to be highest for rural areas.

**Figure 2:**
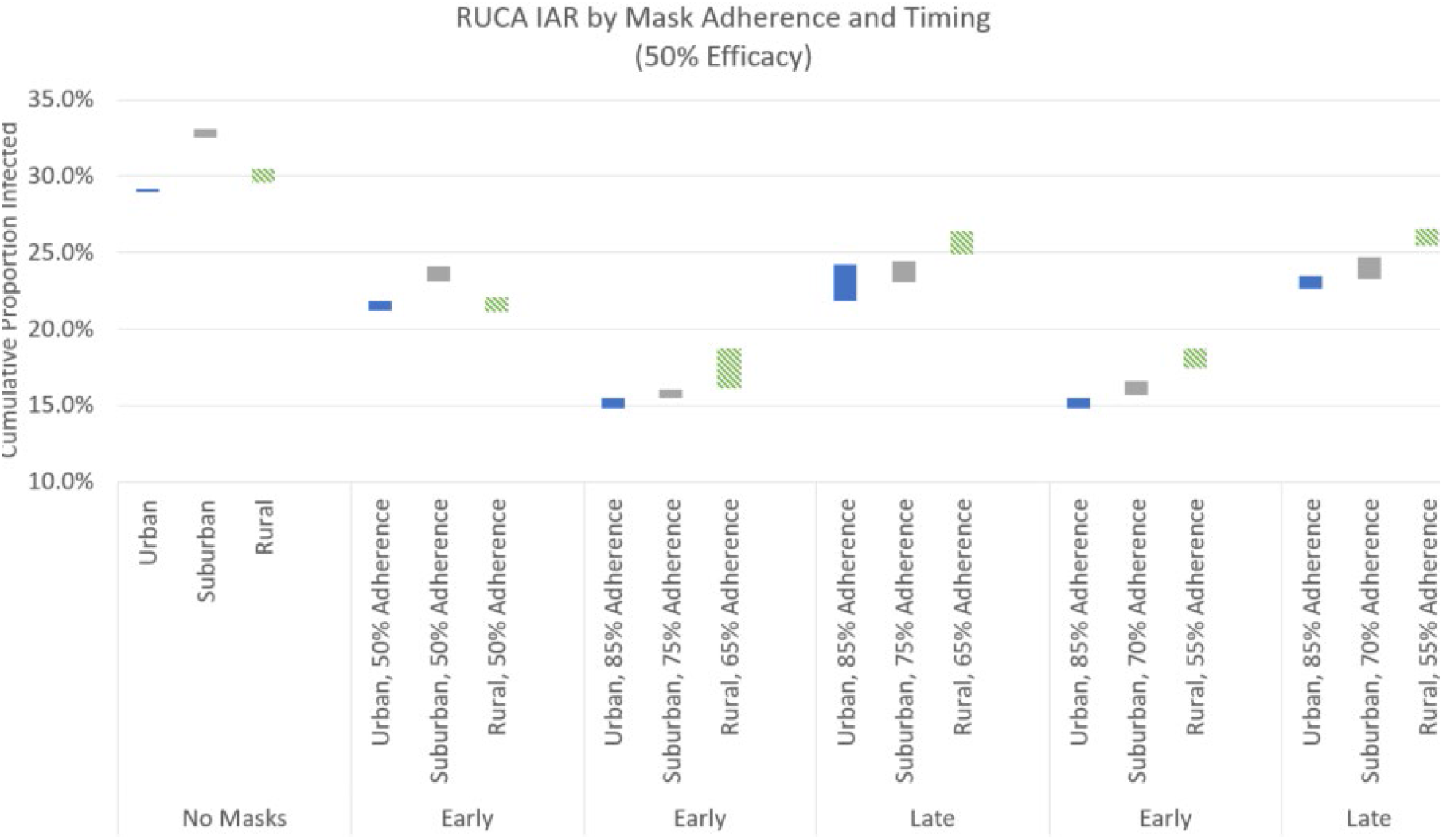
Infection Fatality Rates for areas categorized by urban, rural, and suburban status, where the band is ± 2 standard deviations around the daily mean for all areas in that category across all replications. Mask adherence is shown for 0% adherence, 50% across all areas with early adoption, and two scenarios of differing adherence levels by early and late timing.

Examination of results across demographics indicate that Whites have the lowest IAR and Blacks have the highest mortality rate, findings that are consistent across all masking scenarios.

Finally, the impact of masks on simulation outcomes is additive to the effects of changes in mobility and voluntary quarantining. The control with no masks and no interventions had an IAR of 58%, peak prevalence of 4.7%, 49,000 hospitalizations, and more than 27,000 deaths, while the scenario with mobility changes but 0% mask adherence resulted in 29.6% IAR, 0.74% peak prevalence, 8390 hospitalizations, and fewer than 15000 deaths. In comparison, the results with 50% efficacy and 70% adherence or with 50% adherence and 75% efficacy provided additional improvement over no masks, with IAR below 18%, peak prevalence below 0.3%, hospitalizations below 3300, and deaths below 8600.

## DISCUSSION AND CONCLUSIONS

According to this dynamic model of COVID-19 spread, widespread use of masks after community transmission has begun decreases the impact of the pandemic, even when the overall effectiveness of the intervention is less than 50%. This finding is consistent with empirical findings using publicly reported state-based data from April 1 to May 21, 2020 [17] and a deterministic aggregate model of disease spread [18]. Our modeled scenarios suggest 40% or higher reductions in infections and mortality, over and above those resulting from physical distancing interventions in place, for scenarios with adherence 70% or greater, at least 50% efficacy, and early adoption. Even adoption of mask use many months later in the outbreak can lead to 18% or more reductions in IAR and deaths. We stress that since mask use does not reduce disease transmission to zero in any scenario, other non-pharmaceutical (and future pharmaceutical) interventions need to be maintained.

The finding that rural and suburban areas are at risk for high IAR is consistent with the recent spread of COVID-19 well beyond urban areas [1]. It is somewhat surprising that suburban areas are at higher risk in some scenarios, although these locales have relatively high population density compared to rural areas and their mobility changes have been less than in urban areas (see Supplemental Appendix). Differences in timing of mask adoption and in adherence across geographical areas may eventually be associated with much higher IAR in rural areas that adopted masks late compared to urban ones that adopted early, which was as much as 10 percentage points in the IAR difference in some scenarios from Figure 2. Our demographic results highlight continued disparities for Blacks even under mask mandates, and our results likely underestimate the true demographic gap in infections or mortality.

Our findings lend further support to states or other localities considering mask orders and evidence that such practices have been effective. Improving the quality of masks worn also has the potential to improve population health, e.g., by shifting people from coverings like neck gaiters or bandanas with lower efficacy to masks with multiple fabric layers or special filtration material like N95 masks. Public health organizations should consider adding this to messaging around face masks. In our discussions with health departments, the ability to quantify the improvement from increased mask adoption is also valuable in targeting messaging to the population.

Limitations of this modeling study include potential mischaracterization of SARS-CoV-2 transmission and illness-causing mechanisms, as well lingering data deficits regarding age-based hospitalizations and mortality rates. Some of the key assumptions that are critical include the efficacy of masks and the adherence in the population. The survey data used captures whether people report wearing masks “most or all of the time”, which is insufficient as a true measure of adherence. We do not account for bias in who is wearing a mask, in terms of their social networks or their mobility patterns. We do not incorporate behavioral changes with respect to rising infections; our review of the recent mobility and infection data did not support this consistently across all areas and time periods (unlike earlier analysis by [19]). If this assumption is wrong then we overestimate the impact of masks. If crowded hospitals lead to an increased fatality rate then we underestimate the value of masks in reducing COVID-19-related mortality. The results do not account for vaccination, which we study in a companion paper [7].

In future research, it may be useful to examine the impact of potential covariance of of likelihood of vaccine uptake and likelihood of mask use, and consequent implications for health equity across diverse affected populations.

## CONCLUSION

This simulation of the impact of mask use to counter ongoing COVID-19 pandemic outbreaksprovides evidence that transmission of the SARS-CoV-2 virus can be meaningfully reduced if many people wear masks, especially masks of higher quality. The effect is greater at higher levels of adherence and with masks of higher efficacy, but still meaningful at the lowest levels of each examined. Rural areas with low mask adherence are at particularly high risk for negative outcomes. Specific quantitative values of 20% benefit (late adoption) or 40% (early adoption) may help improve messaging.

## Supporting information

Supplemental material with model details

## Data Availability

The US Census data that was used is publicly available. The paper also uses disease estimates from the literature. Data was obtained from SafeGraph (SafeGraph.com) with the company's permission; similar data can be obtained by other researchers upon request to the company. The data sources are clearly outlined in a supplemental file made available to readers.

## ACKNOWLEDGEMENTS

The project described was supported in part by the National Center for Advancing Translational Sciences (NCATS), National Institutes of Health, through Grant Award Number UL1TR002489. The content is solely the responsibility of the authors and does not necessarily represent the official views of the NIH. The research was also supported by the Council of State and Territorial Epidemiologists and the Centers for Disease Control and Prevention, NC State University, the Fitts Department of Industrial and Systems Engineering at NC State, the Georgia Institute of Technology, and the Cornell Institute for Disease and Disaster Preparedness. The funding agreement ensured the authors’ independence in designing the study, interpreting the data, writing, and publishing the report. Several of the authors are employed by the state universities associated with the sponsorship. We would like to acknowledge Dr. Nicoleta Serban of Georgia Tech for her work on a previous Covid-19 simulation and Dr. Mehul Patel for his leadership on the CovSim initiative.

