## Supplemental material with model details for "High-Quality Masks Reduce COVID-19 Infections and Deaths in the US"

In this document we provide additional information on the model used to project the spread of Covid-19 disease. The model was based on the structures of ones previously published for influenza pandemics [1-3] and for Covid-19 [4]. The model is also similar to that used in [5], where we study vaccine distribution. Portions of the supplemental appendix are the same as in our other papers.

#### Agent-Based Network

Figure S1 summarizes the structure of agents across the network. Data values to generate individual agents were drawn from the US Decennial Census at the census tract level including age in five categories (age 0 to 4; age 5 to 9; age 10 to 19; age 20 to 64; age 65 and greater) [6]. The age groups allow differences with respect to school peer groups (e.g., pre-school, elementary, middle and high school). A household was determined to have children present or not [6, 7], and agents were assigned to households according to the distribution of household size in the tract [6].

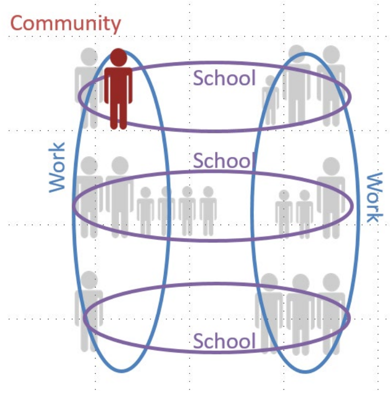


Figure S1: Network of agents allows for interactions and transmission in households, peer groups such as work or school, and communities.

A household was assigned a race/ethnicity according to the population in census tract data (aggregated in the results to Black Only, White Only, Hispanic not Black or White, and Other). Using the publicly available Census data, we assume that race/ethnicity is independent of the other household data such as size. This implies our model is capturing less of the heterogeneity than exists in reality, although comparisons of the outputted data to population estimates show that some aspects are captured due to the similarities within and relatively small size of census tracts. As an indicator for health conditions that could put an individual at higher risk of hospitalization (and subsequently death) due to Covid-19, we used statewide prevalence of diabetes by race/ethnicity [8] adjusted by age. Diabetes affects a large proportion in NC (e.g., 7.8% of Hispanics, 11.4% of Whites, and 14.2% of Blacks) but is only one factor of several high-risk health conditions, so we underestimate the true differences that may exist across the population.

All agents interacted with households at night. During the day, agents age 5 to 19 were allowed interactions in peer groups ("schools", potentially separately for elementary school), and agents 20 to 64 interacted in peer groups ("work") according to commuting patterns. The latter was determined from workflow data [9] indicating the people who live in a census tract and the percentage who work in each of the other census tracts. The model used more than one million agents to represent a state-wide population approximately ten times that size.

For adult agents age 20 to 64, working from home or outside of the house was determined by time-varying mobility factors. Specifically, we used mobility data generated by SafeGraph [10], a data

company that aggregates anonymized location data from numerous applications in order to provide insights about physical places, via the Placekey Community [11]. To enhance privacy, SafeGraph excludes census block group information if fewer than five devices visited an establishment in a month from a given census block group. The SafeGraph data tracks device location data and classifies each as working full time if that device spent greater than 6 hours at a location other than their home geohash-7 during the period of 8 am - 6 pm in local time. The data indicates the count of devices classified as working full-time. As in the publicly available Google mobility data [12], we computed the ratio of devices at home in comparison to the month of January to quantify the proportion of working-age adults staying home. Beginning in month 6 we assume that the mobility factors stay the same. Based on the mobility data from SafeGraph and Google on mobility in contexts outside of work, we assume that the community mobility follows the same pattern as the workplace but to a lesser extent. E.g., If the mobility is at 60% then the community factor is 88% of the original community value (specific calculation for new Community parameter: (1-((1-0.6)/3.5) * (community_parameter = 0.23) = 0.204, where 3.5 is the parameter that was determined by experimentation and sets the scaling with workforce mobility). A reduction in the number of people in the community is modeled by a reduction in the community infectious hazard. In the workplace, the reduction of people follows the workplace mobility values. This decrease in workplace activity reduces the workplace infectious hazard. Because of sparseness of data in rural areas, we grouped SafeGraph data at the census tract level stratified by urban, rural, or suburban according to the RUCA classification system and whether the tract was in the 1st, 2nd, 3rd, or 4th quartile of household median income for the state level. To validate the approach, we compared to the Google mobility data, which is available at the county level [12]. Examples of mobility factors from the analysis are included below.

For voluntary quarantining, we assume 30% of households do this initially, then 60% beginning day 46 to correspond to increased messaging. As in Keskinocak et al. (4) that validated values computationally, the value decreases by 0.15 points per week until reaching 15%, where it remains.

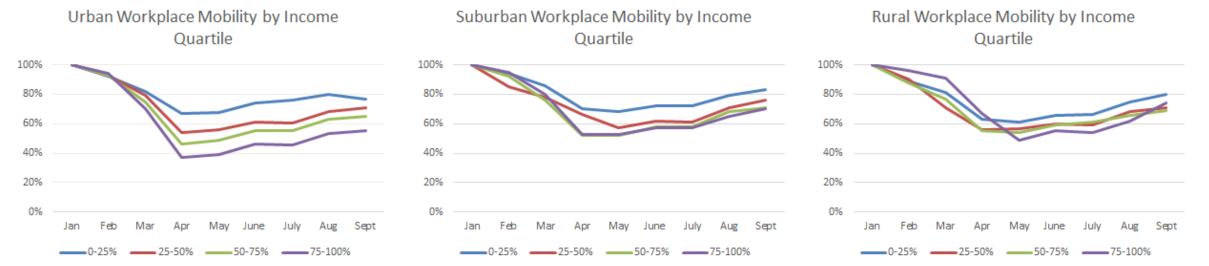


Figure S2: Mobility over time stratified by urbanicity and median household income level (four quartiles) of the area

### Disease Progression within an Individual

The underlying model of disease spread was assumed to be a variant of a Susceptible-Exposed-Infectious-Recovered (SEIR) model; see Figure S3. At a given point in time, each individual is in exactly one state: susceptible (S), exposed (E), pre-symptomatic (IP), asymptomatic (IA), symptomatic (IS), hospitalized (H), recovered (R), or dead (D). In this paper, we aggregate all hospitalizations regardless of use of Intensive Care Unit or ventilator.

The full details of the model are available in Keskinocak et al [4] in the supplemental document. A portion of the material in that document is summarized in this subsection.

The states in the disease progression model include Susceptible (S), Exposed (E), Infectious Pre-symptomatic (IP), Infectious Asymptomatic (IA), Infectious Symptomatic (IS), Hospitalized (H), Recovered (R), and Dead (D). An individual is in exactly one state at a time, and all individuals begin as susceptible. Movement from Susceptible to Exposed depends on contact with an infected person, infectivity of the other contact, and susceptibility of the initial person, given any interventions in place. From Exposed, an individual would move to IP, and from there to either IA or IS. From IS an individual can move to Hospitalized or Recovered, and from Hospitalized to Death or Recovered.

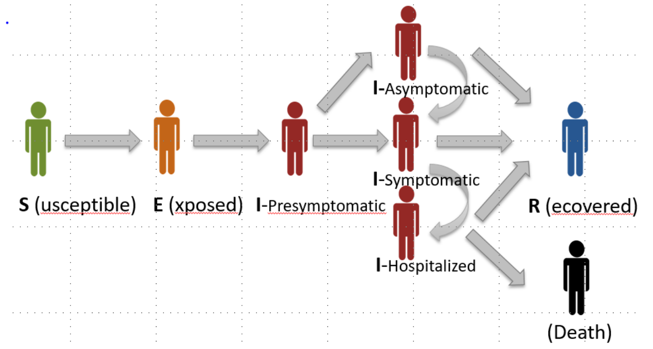

Figure S3: SEIR-type model represents the progression of Covid-19 for an individual.

The list of input parameters and associated references is provided in Table S1. Additional details are available in the supplement of Keskinocak et al. [4].

Several parameters determine the length of time within each state in the SEIR including the mean and standard deviation on the time before an exposed patient becomes pre-symptomatic, the average length of time of the pre-symptomatic phase, the distribution of time within the symptomatic (S) stage, the distribution of the length of hospitalization, and the ratio of the duration of the symptomatic and asymptomatic states. Parameters related to the transmission between states include the probability of moving to symptomatic (from IP), the probability of hospitalization (from IS), and the probability of death (from H); probabilities out of a state (including to itself) must sum to 100%. The probability of hospitalization is specific to whether someone has diabetes or not, matching both the relative risk [13, 14] and the overall hospitalization rate. Note that in our model death only occurs after a hospitalization. In reality there are some cases where death occurs absent a hospitalization; this implies that our hospitalizations may be an overcount of the true ones. One could also think of our hospitalized state as having a case severe enough that hospitalization is needed.

The overall infection fatality rate (IFR) (i.e., the probability that an exposed person will die) in our model relates to both the probability of hospitalization and the probability of death from that state. In baseline runs of the simulation, the computed IFR across all age groups is 0.4% to 0.5%. This is consistent with the lowest country values reported in [15] and the lowest regional values in the US from [16].

The infectivity of the virus at the beginning of the outbreak without interventions is summarized by reproductive rate R0, and the transmission rate (denoted as $\beta$). The proportion of transmissions that occur at either the IP or IA stage is $\theta$, and the proportion of infections generated by individuals who are never symptomatic is $\omega$. In the absence of interventions, the proportion of transmission that occurs outside households is $\gamma$, and the proportion of transmission outside households that occur in the community is $\delta$.

One of the important differences in comparison to the values used for influenza [1], is that the proportion of transmissions that can occur by people without symptoms is much higher. In comparison to the Covid-19 values used for the state of Georgia [4], $\delta$ is a little lower, reflecting the spread in NC calibrated computationally. For this paper we also take the import rate of cases to be lower (45 compared to 100) based on factors such as the airport size, commuting in or out of the state, etc. Like [4], our hospitalization rates (which are conditional on symptomatic cases) were lowered in model calibration.

**Validation**

The simulation was calibrated and validated on cumulative cases [17], deaths [17], and hospitalizations [18]. Deaths and hospitalizations are appropriate for direct validation; while not perfect, they are a reasonable estimate of true hospitalizations and deaths. However, it is well-recognized that the true cases are much greater than those that are lab-confirmed, and this value has changed over time. In July, the CDC published estimates that the true cases had been approximately 10 times the lab-confirmed cases [19], and later published estimates that the rate was still as much as eight times higher in some locations [20]. Using the Infection Fatality Rate (IFR) of 0.5 [15, 16] and the average time from exposure to death of 21 days (consistent with [21]), we computed the daily expected number of true cases that would have been needed to result in the recorded deaths. The average daily ratio of computed cases to lab-confirmed cases (or the "lab-multiplier") ranges from approximately 10 initially to 4 (after 147 days). The estimates obtained in this way for one state and nationally are also consistent with CDC published accounts of antibody testing to identify the number of true cases [19, 20]. This approach does not take into account the changing age-distribution of positive cases, which likely affected the timing of deaths. We compare our simulated results to actual deaths and hospitalizations and cases adjusted by the lab multiplier. This adjustment is only for validation, as the simulation reports the projected cases and deaths (not lab-adjusted cases). For the baseline scenario, our estimates are within 6% of the reported cumulative values after more than 200 days without updating cases after March 24.

**Stochastic Simulation across Network**

To seed infections in agents, the confirmed cases at the county level over time were collected from data hosted by The New York Times (22). Infections were assigned to census tracts randomly according to the population within each tract in the county using the Huntington-Hill method. For validation, data was also collected on deaths (The New York Times) and hospitalizations (23).

For each scenario, 15 replications of the simulation are run. This balances the computational time with the variability in results generated across replications. The specific number of replications needed would also vary according to what model parameters are of interest (e.g., more would be needed to reduce uncertainty in results at the county level). Results were checked for convergence. A given replication begins with a random seed, which leads to several realizations of that randomness for the replication: (i) the specific network, e.g., the random assignment of a particular agent and their attributes (household size, commuting location, diabetes or not, etc.) as described in the model above; (ii) the individual agents who are infected with the seed infections; (iii) whether an infected individual will transmit the virus to another person in the household, peer group, and/or community at a point in time; and (iv) the duration within a disease state. The mean and standard deviation of the performance metrics are computed across the simulation replications. Figures in the paper with measures over time also report the band associated with plus or minus two standard deviations around the mean.

Table S1: Displays key model parameters and references. Many parameters are also summarized in the supplemental material of Keskinocak et al [4].

| PARAMETER | ESTIMATES | REFERENCES |
|---|---|---|
| Exposed (E) Duration | Weibull with mean 4.6 days | [22, 23] |
| Pre-symptomatic (IP) Duration | 0.5 days | [22] |
| Hospitalized (H) Duration | Exponential with mean 10.4 days | [22, 24] |
| Symptomatic (S) Duration | Exponential with mean 2.9 days | [15] |
| Symptomatic-Asymptomatic Duration Ratio | 1.5 | [22] |
| Probability of Symptomatic (from IP) | 0.50-0.82 | [25]<br>[26-28] |
| Q1` | 0.016 for age 0-19;<br>0.056 for age 20-64;<br>.08 for age 65+ | (17), [29, 30], and calibration |
| Probability of Hospitalization for diabetes | 3 times above rate | [13] |
| Probability of Death (from H) | 0 for age 0-19;<br>0.0515 for age 20-64;<br>0.3512 for age 65+ | [31] |
| R0 | 2.4 | [32-34] |
| $\beta$ transmission rate | 1.12 | [32] |
| $\theta$ (probability IP to IA) | 0.48 | [35] |
| $\omega$ (proportion infections by IA) | 0.24 | [35] |
| $\gamma$ (proportion of transmission that occur outside households) | 30% | [1, 4] |
| $\delta$ (proportion of infections outside households that occur in community) | 0.23 | [4]; Calibration |
| FlatImportRate | 45 | [4]; Calibration |
